# Viral Variants and Vaccinations: If We Can Change the COVID-19 Vaccine… Should We?

**DOI:** 10.1101/2021.01.05.21249255

**Authors:** Sharon Bewick

## Abstract

As we close in on one year since the COVID-19 pandemic began, hope has been placed on bringing the virus under control through mass administration of recently developed vaccines. Unfortunately, newly emerged, fast-spreading strains of COVID-19 threaten to undermine progress by interfering with vaccine efficacy. While a long-term solution to this challenge would be to develop vaccines that simultaneously target multiple different COVID-19 variants, this approach faces both developmental and regulatory hurdles. A simpler option would be to switch the target of the current vaccine to better match the newest viral variant. I use a stochastic simulation to determine when it is better to target a newly emerged viral variant and when it is better to target the dominant but potentially less transmissible strain. My simulation results suggest that it is almost always better to target the faster spreading strain, even when the initial prevalence of this variant is much lower. In scenarios where targeting the slower spreading variant is best, all vaccination strategies perform relatively well, meaning that the choice of vaccination strategy has a small effect on public health outcomes. In scenarios where targeting the faster spreading variant is best, use of vaccines against the faster spreading viral variant can save many lives. My results provide ‘rule of thumb’ guidance for those making critical decisions about vaccine formulation over the coming months.

## Introduction

On March 11, 2020, the World Health Organization (WHO) declared the COVID-19 outbreak a global pandemic^1^. Since then, much effort has focused on developing safe and effective vaccines that can bring the world back to normality^2^. Remarkably, exactly 9 months after the WHO declared a global pandemic, the U.S. Food and Drug Administration (FDA) announced an Emergency Use Authorization (EUA) for the Pfizer-BioNTech COVID-19 vaccine^3^ – the first vaccine against COVID-19 to receive approval in the United States. Additional authorizations have followed rapidly. For instance, an EUA was issued for the Moderna COVID-19 vaccine a mere one week after the EUA for the Pfizer-BioNTech vaccine^4^. Notably, this timeline for vaccine development and approval is over four times faster than any previous vaccine efforts^5^. Further, initial clinical trials suggest that these early vaccines against COVID-19 are highly effective, with both Pfizer-BioNtech and Moderna reporting efficacies exceeding 90%^6,7^, which is comparable to some of the best vaccines currently in use.

In large part, the remarkable speed with which the world has developed safe, efficacious COVID-19 vaccines is a result of years of advances in molecular biology. Indeed, both the Pfizer-BioNTech vaccine and the Moderna vaccine rely on a novel messenger RNA (mRNA) approach^8,9^ for stimulating the immune system. Though there are some drawbacks of mRNA vaccines – most notably the requirement for cold-chain storage – this technology greatly facilitates rapid and targeted vaccine development based solely on the genetic sequence of the virus^10^. This, in turn, allows vaccine candidates to be made without ever needing to culture the virus itself. Another related advantage of mRNA vaccines is their flexibility. In particular, because their only requirement is the genetic code for a particular antigenic component of the target virus, it is not difficult to alter these vaccines in response to strain variation.

In early December 2020, the United Kingdom reported a novel and potentially concerning COVID-19 variant, B.1.1.7^11,12^. What is particularly troublesome about this variant is that it has a large number of mutations – 23 in total, 17 of which are non-silent^12^. Further, 8 of these mutations occur in the spike protein, which is the portion of the virus responsible for enabling viral entry into human cells^12^. These novel mutations include the N501Y mutation that allows the virus to bind more tightly to the human angiotensin converting enzyme 2 (ACE2) receptor^13^, as well as the D614G deletion that appears to make the virus more transmissible between individuals^14^. Substantial genetic changes, along with accelerating rates of spread, and a rapid rise to dominance of the B.1.1.7 variant in Southern England^15^, suggest that the B.1.1.7 has a fitness advantage. In fact, recent modeling studies indicate that the B.1.1.7 variant could be 50-70% more transmissible than previous COVID-19 strains^11,16^.

Beyond altered transmissibility, another threat of the B.1.1.7 mutant is that existing vaccines may be less effective against it. This is because most vaccines in development specifically target the viral spike protein^2^. Currently Pfizer-BioNTech and Moderna are conducting in vitro assays that will provide first estimates of the efficacy of existing vaccines against the novel B.1.1.7 COVID-19 variant. Full assessment of vaccine efficacy will follow in a matter of weeks to months, as existing vaccines are rolled-out in locations like Southern England where the B.1.1.7 variant is dominant. However, even if current vaccines are efficacious against B.1.1.7, the emergence of this viral variant is a warning call. In particular, we can expect COVID-19 to continue to mutate and to generate novel variants that differ from the initial vaccine targets. The obvious long-term solution is to develop vaccines that contain DNA from all of the dominant COVID-19 variants in circulation (multivalent), much like the seasonal influenza vaccine^17^. However, because mRNA vaccine technology is new, multivalent COVID-19 mRNA vaccines have not been tested. Thus, it is unclear whether presenting multiple antigenic components through mixed mRNAs will actually provide even immune coverage to all components, or whether there will be interference effects^18^. Likewise, it is unclear whether there are any additional health risks to mixed mRNA vaccines. As a consequence, multivalent COVID-19 vaccines pose both development and regulatory hurdles. A simpler approach to combat COVID-19 strain evolution is to ‘switch out’ the viral mRNA in the existing single strain (monovalent) vaccine in response to changes in the dominance of the circulating strains. Currently, for example, that could mean switching to the mRNA for the spike protein from the B.1.1.7 viral variant.

While it may seem obvious that COVID-19 vaccines should be altered to track the dominant viral strains in circulation, there are costs involved with switching. Further, without sufficient cross-protection, vaccination against one viral variant will cause the other to rise to dominance and vice versa, essentially leading to a game of whack-a-mole. While this scenario cannot be avoided entirely, there may still be better and worse decisions in terms of the viral variant that is selected for vaccine efforts. This is because not all viral variants are biologically equivalent. Some viral variants, like B.1.1.7 are more transmissible. Others may be more prevalent, or may have already swept a larger fraction of the population, at least at the time of the initial vaccine roll-out. This, then, begs the question: When multiple COVID-19 viral variants are circulating, and when vaccine cross-protection is not complete, which variant should be targeted for vaccine scale-up during the initial vaccination phase?

In this paper, I use a simple stochastic simulation to examine the outcomes of various vaccine strategies against two different co-circulating COVID-19 viral variants. In particular, I examine how the best choice of vaccine target depends on the degree of cross-protection offered by vaccination and/or natural immunity to the alternate strain. I also consider how outcomes depend on the relative transmissibilities of the different viral variants, their relative prevalence at the beginning of the vaccination period, and existing natural immunity. Finally, I consider how differences in the timing of vaccine roll-out can change predictions, and what this might mean as we rapidly scale-up a series of different mRNA vaccines against the COVID-19 pandemic.

## Method

I use the Gillespie algorithm (GA)^19^– a discrete-time, event-based simulation approach – to study transmission of two different COVID-19 viral variants in a human population undergoing rapid roll-out of a COVID-19 vaccine that is targeted at only one of the two variants (monovalent). For each individual in the population, I assume that they can be in one of four potential infection classes, one of four potential natural immunity classes and one of four potential vaccination classes. Specifically, individuals can be: virus-free (*i* = 0), infected with the first viral variant (*i* = 1), infected with the second viral variant (*i* = 2) or infected with both viral variants (*i* = 3). Likewise, individuals can be: fully naturally susceptible (*j* = 0), naturally immune to the first viral variant (*j* = 1), naturally immune to the second viral variant (*j* = 2) or naturally immune to both viral variants (*j* = 3). Finally, individuals can be: unvaccinated (*k* = 0), vaccinated against the first viral variant (*k* = 1), vaccinated against the second viral variant (*k* = 2) or vaccinated against both viral variants (*k* = 3). Ultimately, this leads to 4×4×4 = 64 possible states for each individual. I then define a matrix, *N*, where each element, *N*_*i,j,k*_, specifies the number of individuals in the *i*^th^ infection class, the *j*^th^ natural immunity class and the *k*^th^ vaccination class. Depending on model assumptions, not all states may be possible, in which case *N*_*i,j,k*_ = 0 (e.g., if there are no double vaccinations, then *N*_*i,j*,3_ = 0).

### Viral Transmission

I assume frequency-dependent viral transmission. Thus, the probability of a new viral infection involving viral variant *i* is proportional to both the number of individuals infected with that variant and the number of individuals susceptible to that variant, and is inversely proportional to total population size. I assume that natural immunity and/or vaccine-induced immunity impact the probability of transmission of each viral variant, but are not necessarily fully protective, even against the ‘on-target’ viral variant. Specifically

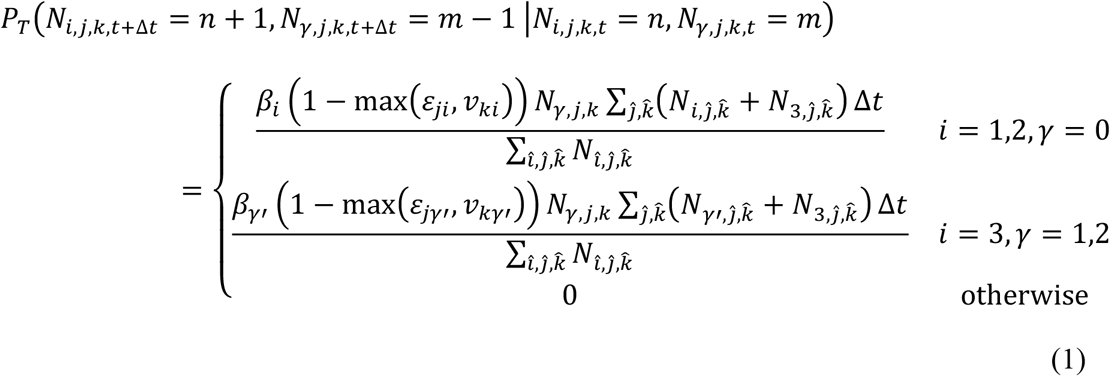

where *P*_*T*_(*N*_*i,j,k,t+*Δ*t*_=*n*+1, *N*_*γ,j,k*,t+Δ*t*_ = *m* – 1 | *N*_*i,j,k,t*_ = *n, N*_*γ,j,k*,t+Δ*t*_ = *m*) is the probability of an individual transferring from the A infection class to the *i* infection class and 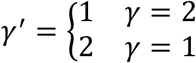.

In equation (1), *β*_*i*_ is a parameter that describes the transmissibility of viral variant *i, ε*_*ji*_ is a parameter that describes the reduction in transmission of viral variant *i* to an individual of immunity state *j* and *v*_*ki*_ is a parameter that describes the reduction in transmission of viral variant *i* to an individual of vaccination state *k*. For all models that I consider, I assume that *ε*_*ji*_ = *v*_*ki*_ when *j* = *k* (natural immunity and vaccine induced immunity confer similar levels of protection). Further, I assume that *ε*_11_ = *ε*_22_ = *v*_11_ = *v*_22_ and that *ε*_12_ = *ε*_21_ = *v*_12_ = *v*_21_ (both viral variants perform similarly in terms of the degree of conferred same-strain protection and the degree of conferred cross-strain protection). Notice that equation (1) makes several simplifying assumptions. First, a current infection with one viral variant does not impact the likelihood of a secondary infection with the alternate viral variant. Second, co-infected individuals spread each viral variant at the same rate as if they were singly infected. Third, immunity and vaccination status do not impact the extent to which an infected individual spreads disease. Fourth, co-infections are picked up serially, rather than as a result of direct transfer from a co-infected individual. All of these assumptions could be relaxed, though with the requirement for additional parameters.

### Recovery

I assume a constant probability of recovery of infected individuals. Further, I assume that, upon recovery, an individual acquires natural immunity to the viral variant that they were infected with (or remains naturally immune, if they already had immunity to that particular strain). Thus,

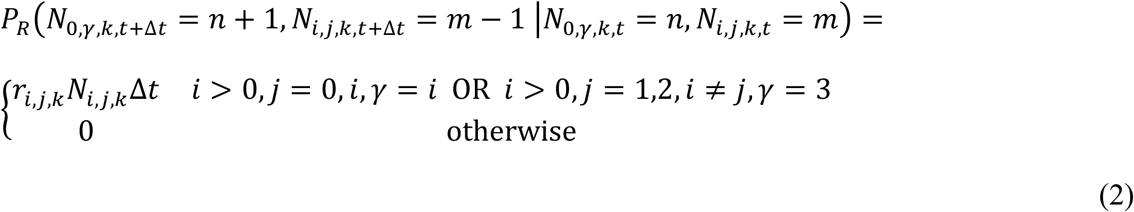

where *P*_*R*_(*N*_*i,j,k,t+*Δ*t*_= *n* + 1, *N*_*γ,j,k*,t+Δ*t*_ = *m* – 1 | *N*_*i,j,k,t*_ = *n, N*_*γ,j,k,t*+Δ*t*_ = *m*) is the probability of an individual transitioning from the *i* infection class to the virus-free infection class (*i =* 0) and the A immunity class. In equation (2), the rate of recovery can depend on the viral variant, on whether an individual is infected with a single viral variant or is co-infected, and on the immunity and vaccination status of the infected individual. However, in all models that I consider, I assume *r*_*i,j,k*_ = *r*; thus all individuals have similar probabilities of recovery, regardless of disease state or immunity status.

### Death

I assume a constant probability of death of infected individuals and do not consider additional death or birth in the population. Specifically

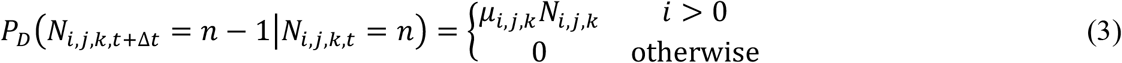

where *P*_*D*_ (*N*_*i,j,k,t +*Δ*t*_ = *n* –| *N*_*i,j,k,t*_ = *n*) is the probability of an individual from the *i* infection class being removed from the population. In equation (3), the probability of dying depends on the particular viral infection, as well as the immunity status of the host. However, as with recovery, for all models that I consider, I assume that *μ*_*i,j,k*_ = *μ*_*i*_. This means that death rate is only a function of the viral variant(s) causing the infection but does not depend on immune status of the host.

### Vaccination

I assume a constant probability of vaccination, and that vaccination does not take into account the natural immunity status or infection status of the individual. I make this assumption given the large number of asymptomatic infections that make it difficult to know whether a person has been previously infected or, indeed, even whether a person is currently infected. I do, however, assume that people who have already been vaccinated are not re-vaccinated. Finally, I assume that vaccination proceeds at a fixed rate until all individuals willing to receive a vaccine have done so. Once all willing individuals have been vaccinated, vaccination stops. Thus

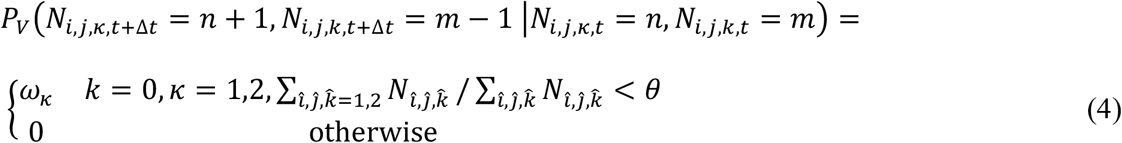

where *P*_*V*_ (*N*_*i,j,k,t +*Δ*t*_ = *n* + 1, *N*_*γ,j,k,t*+Δ*t*_ = *m* – 1 | *N*_*i,j,k,t*_ = *n, N*_*γ,j,k,t*_ = *m*) is the probability of an individual with vaccination status M receiving the vaccine and transitioning to vaccination status *κ, ω*_*κ*_ is the rate at which the vaccine is administered and *θ* is the fraction of the population willing to receive a vaccine.

### Parameters and Initial Conditions

Except where noted otherwise, I assume the parameter values as outlined in Table 1. Briefly, I assume that both vaccines and natural immunity are 95% effective at preventing infection when targeted at the same viral variant. This is commensurate with early reports from both Pfizer-BioNTech and Moderna^6,7^, as well as low reports of re-infection across the world. Likewise, I assume that a person who has been infected can spread the virus for 10 days^20^ (notice that it does not matter whether the person has symptoms for any or all of this period). I assume that the death rate due to disease is 0.0006/day which, for a 10 day infection period, leads to a 0.6% death rate, again commensurate with current reports on COVID-19 infection fatality ratios (IFRs)^21^. Finally, I assume that a person infected with the original COVID-19 variant spreads the disease to, on average, 2 additional people^22^, while a person infected with the new viral variant spreads the disease to, on average, 3 additional people (i.e., 50% more infectious). Finally, I assume that 100% of the population is willing to/forced to receive the vaccine, and that 5000 people per day, can be vaccinated in a 500,000 person population (i.e., it takes 100 days or approximately 3 months to vaccinate the entire focal population).

**Table 1:** Parameters and initial conditions used in simulations

| Parameter | Symbol | Value |
| --- | --- | --- |
| Natural immunity against same variant | $\varepsilon_{11}, \varepsilon_{22}$ | 0.95<sup>6,7</sup> |
| Natural cross-protection against the opposite variant | $\varepsilon_{12}, \varepsilon_{21}$ | 0.70 |
| Vaccine induced immunity against same variant | $v_{11}, v_{22}$ | 0.95 |
| Vaccine induced cross-protection against the opposite variant | $v_{12}, v_{21}$ | 0.70 |
| Recovery rate | $r$ | 0.1<sup>20</sup> |
| Death rate | $\mu_1, \mu_2$ | 0.0006<sup>21</sup> |
| Transmission rate of COVID-19 variant 1 | $\beta_1$ | 0.2<sup>22</sup> |
| Transmission rate of COVID-19 variant 2 | $\beta_2$ | 0.3 |
| Fraction of population willing to receive vaccine | $\theta$ | 1 |
| Vaccination rate | $\omega_1 + \omega_2$ | 5000 |
| Population size | $\sum_{i,j,\hat{k}} N_{i,j,\hat{k}}$ | 500000 |
| Initial fraction infected with variant 1 | $\rho_1$ | 0.001 |
| Initial fraction infected with variant 2 | $\rho_2$ | 0.00001 |
| Initial fraction naturally immune to variant 1 | $\varphi_1$ | 0.10 |
| Initial fraction naturally immune to variant 2 | $\varphi_2$ | 0 |

For initial conditions, I assume that, at the start of the simulation, a pre-defined fraction of the population, *ρ*_1_, is infected with the first viral variant, and a pre-defined fraction of the population, *ρ*_2_, is infected with the second viral variant. I ignore co-infections at the beginning of the simulation under the assumption that 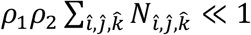 for reasonably small *ρ*_1_ and/or *ρ*_2_. Likewise, I assume that, at the start of the simulation, a pre-defined fraction of the population, *φ*_1_, is immune as a result of previous infection with the first viral variant, and that a pre-defined fraction of the population, *φ*_2_, is immune as a result of previous infection with the second viral variant. As with infection, I assume that there are no individuals who have acquired natural immunity to both viruses, which is a good approximation when either *φ*_1_ or *φ*_2_ are small (at least one strain has not been circulating for a long time) or when *ε*_12_ or *ε*_21_ are large (natural cross-protection is high, making serial infection with different strains less likely). In practice, most simulations that I consider assume that the second COVID-19 strain is a recent introduction, and thus *φ*_2_∼0.

Python code for model simulations is provided at: https://github.com/bewicklab/COVID-Vaccination-Strategies

## Results

In the analysis that follows, I consider three different vaccination strategies. First, I consider a scenario where all doses of the vaccine are targeted against viral variant 1 (blue lines). Second, I consider a scenario where all doses of the vaccine are targeted against viral variant 2 (red lines). Finally, I consider a scenario where two different types of vaccines are in use, with 50% of the doses targeted against viral variant 1, and 50% of the doses targeted against viral variant 2 (purple lines; this could be achieved, for example, if Pfizer-BioNTech and Moderna produced different vaccines, each targeting the opposite viral variant). For each scenario, I consider three metrics: (1) total deaths from the start of the simulation until the virus goes extinct, (2) peak number of infections at any given time and (3) number of days until the virus goes extinct. Total deaths provides an estimate of the costs of the different strategies in terms of human life. Total deaths are also proportional to total infections, thus giving a sense of the overall scale of the viral outbreak for each scenario. Peak number of infections is important, because an outbreak that has a higher number of infectious individuals at any single point in time is more likely to overwhelm healthcare facilities. Thus, even if the total sizes of two different outbreaks are the same, death rates are likely to be lower for the outbreak with a lower peak infection rate (which typically implies an outbreak that is spread over a longer period of time – note that my model assumes a constant death rate, thus total deaths does not account for variation in death rate due to overwhelmed medical facilities). Finally, the number of days until the virus goes extinct gives a sense of the time required to reach herd immunity through a combination of natural infection and vaccination. Longer times to viral extinction do not necessarily mean worse outbreaks in terms of total infections and deaths, although they do imply a longer period over which masks and social distancing may be required, particularly for at-risk people. For all simulations, I assume that the first viral variant spreads at a slower rate (i.e., is less transmissible) than the second.

### Degree of Cross-Protection

Figure 1 shows the total number of deaths (A), peak infections (B) and time to virus extinction (C) as a function of the degree of cross-protection offered by natural immunity and/or vaccination to the opposite viral variant. For these simulations, I assume a total population size of 500,000 individuals, and an IFR of 0.6%. Consequently, if the entire population is infected with one of the two virus strains, then approximately 3000 individuals will die. Likewise, if the entire population is infected with both of virus strains (simultaneously or sequentially), then approximately 6000 individuals will die. Because herd immunity is typically achieved at infection rates less than 100%, 3000 and 6000 deaths represent upper bounds for single infections and double infections respectively. Nevertheless, these bounds help to frame simulation results.

**Fig. 1.**
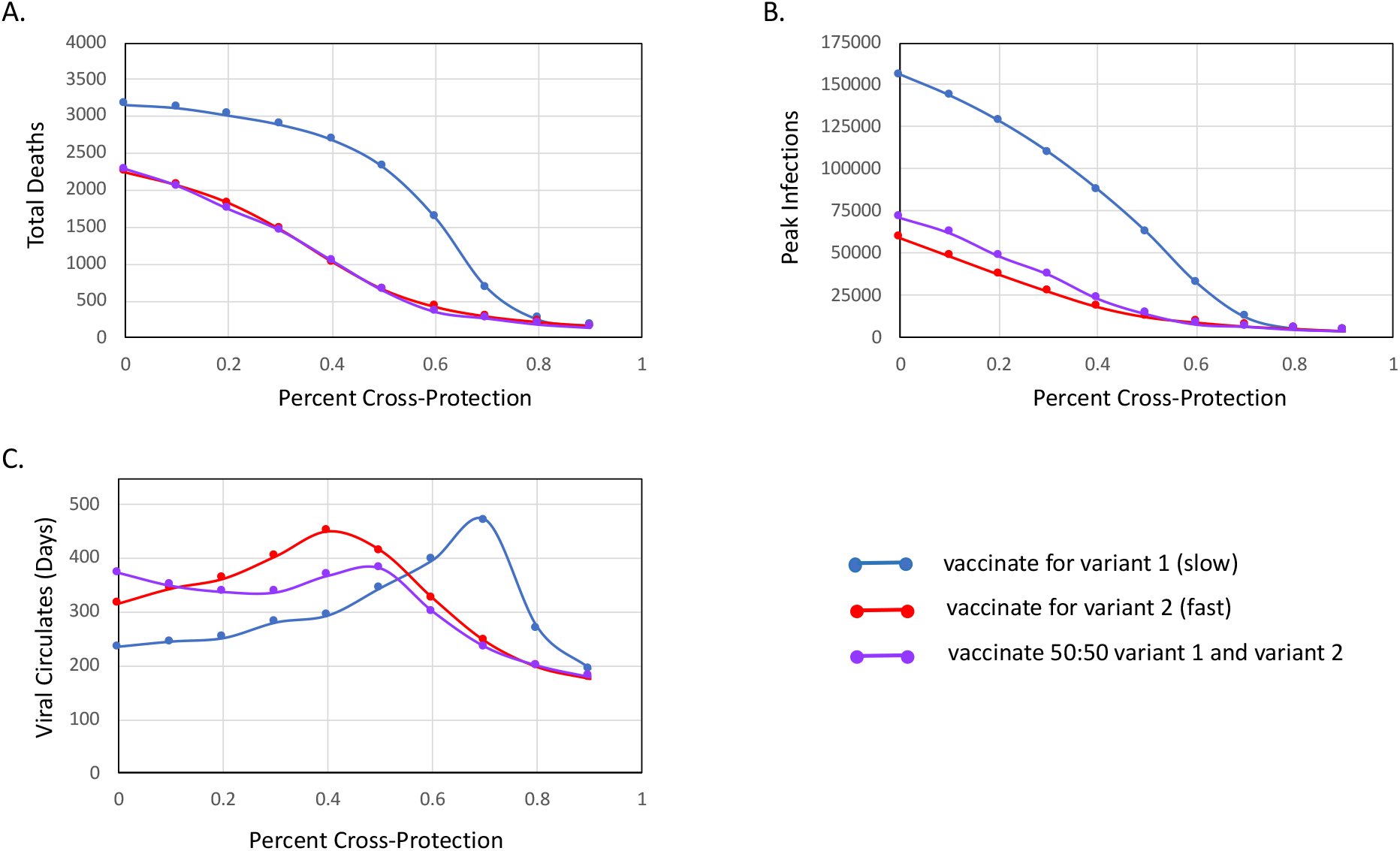
(A) Total number of deaths, (B) peak infections and (C) time to virus extinction as a function of the degree of natural and vaccine-induced cross-protection *v*_12_ = *v*_21_ = *ε*_12_ = *ε*_21_) assuming a vaccination strategy targeting viral variant 1 (blue, slower spreading), a vaccination strategy targeting viral variant 2 (red, faster spreading) and a mixed strategy with 50% of the population receiving the vaccine against viral variant 1 and 50% receiving the vaccine against viral variant 2. All parameters and initial conditions except those associated with cross-protection (*v*_12_, *v*_21_, *ε*_12_, *ε*_21_) are as defined in Table 1. Results shown are median values over 30 simulation trials.

As expected, when cross-protection is close to zero, vaccination against one of the two viral strains cuts the total death rate in half. However, it does nothing to prevent deaths from the off-target viral variant. Because of this, vaccination against the more transmissible virus (in this case, viral variant 2) is almost always optimal, even if the less transmissible virus is initially present at substantially higher rates. This is because, regardless of initial conditions, the off-target variant will inevitably sweep the population. The virus that is more transmissible, however, will infect a higher proportion of the population prior to reaching herd immunity. Consequently, vaccinating against this more transmissible strain is the better option when there is little to no cross-protection.

Although faster spread of viral variant 2 is the primary factor favoring use of this strain as the vaccine target at low levels of cross-protection, there are two additional advantages to focusing on viral variant 2. Both are related to the fact that viral variant 1 is more prevalent. While somewhat counterintuitive, higher prevalence of a particular strain prior to vaccination can actually make it less effective to target that strain with the vaccine (see Fig. 5). This is because higher viral prevalence goes hand-in-hand with higher levels of natural immunity, which has two consequences. First, if one of the two viral variants will inevitably sweep the population (i.e., low cross-protection), then it is preferable that this be the variant with more existing natural immunity, since fewer additional deaths will be necessary to reach herd immunity. Second, when the vaccine is targeted against the more prevalent variant, more vaccine doses are wasted protecting individuals who are already naturally immune (see Figure 5). By contrast, wasted vaccine doses less common when the vaccine protects against a strain without much pre-existing natural immunity. Again, this disfavors use of the more prevalent strain as the vaccine target (in this case, variant 1).

Interestingly, a relatively high level of cross-protection is required before vaccination with the more prevalent but slower spreading variant becomes a competitive strategy. Increasing cross-protection from 0% to 50%, for instance, only reduces total deaths by <30% when the vaccine is targeted against the prevalent but slow spreading variant 1. By contrast, cross-protection has a much stronger effect earlier on when the vaccine is targeted against the fast spreading variant 2. In this case increasing cross-protection from 0% to 50% causes a >70% reduction in deaths. Notably, even at 90% cross-protection, vaccination against the slow spreading variant still underperforms in terms of preventing deaths.

Surprisingly, a mixed strategy, where half of the population receives the vaccine against the slow spreading variant, while the other half receives the vaccine against the fast spreading variant, performs almost as well as fully targeting the fast-spreading variant. This is particularly notable, since a mixed strategy may be more realistic to implement, at least in the near-term, for COVID-19. A mixed strategy, for example, would only require one or a few vaccine-makers to alter the formulations of their vaccines. Notice, however, that my simulations assume randomization of the two vaccines throughout the entire vaccine roll-out period – thus results could be different if the two different vaccines were rolled out on different timelines or in different spatial locations.

Like deaths, peak infections are also much higher under the scenario with vaccination against the prevalent but slow spreading variant 1. Interestingly, however, peak infections are also higher for the mixed vaccination strategy as compared to vaccination against viral variant 2, at least when cross-protection <50%. This contrasts what was seen for total deaths. Nevertheless, the mixed strategy still performs substantially better than vaccination against the slow spreading variant 1 over most of the range of potential cross-protection levels. At high cross-protection, (80-90%), differences in the different vaccination strategies are minimal, and the mixed strategy, or even vaccination against the slow spreading viral variant can actually yield lower peak infection levels.

Times to viral extinction tend to be unimodal, at least for strategies that target a single viral variant. This is because, at low cross-protection, the off-target variant rapidly sweeps the population, driving its own demise over a short period of time (at least in the absence of introduction of newly susceptible individuals through birth or waning immunity). Meanwhile the vaccine rapidly suppresses the on-target variant. Conversely, at high levels of cross-protection, the vaccine itself leads to rapid extinction of both variants. At intermediate levels of cross-protection, however, spread of the off-target viral variant is slowed, but not stopped, leading to longer persistence of this strain in the population. While viral persistence peaks around 40% cross protection for the mixed strategy and the strategy that targets the fast-spreading variant 2, it peaks at a much higher 70% cross protection for the strategy that targets the slow-spreading variant 1. Alarmingly, when the vaccine targets viral variant 1, the virus can persist nearly 1.5 years after vaccination begins, even with a rapid vaccine roll-out of 3 months.

### Relative Transmission Rates

Figure 2 shows the total number of deaths (A), peak infections (B) and time to virus extinction (C) as a function of the transmission rate of the faster spreading viral variant 2. In these simulations, I assume that viral variant 1 spreads at a rate of 0.2 person^-1^day^-1^. For a typical infectious period of 10 days, this means that each person, on average, spreads the virus to two additional people in a fully susceptible population. This estimate is commensurate with current measures of *R*0 for COVID-19^22^. By extension, for the range of transmissibilities considered in Figure 2, each person infected with viral variant 2 transmits the virus to between 2 and 4 additional individuals in a fully susceptible population. This is in keeping with the estimate that the new B.1.1.7 COVID-19 strain is 50-70% more transmissible than its predecessor^11,16^. Not surprisingly, increasing transmissibility of the second viral variant increases overall deaths, as well as peak infections at the height of the outbreak. However, the rate of increase depends on vaccination strategy. When there is not much difference between the transmission rates of the two viral variants, it is actually best to vaccinate against the more prevalent but slower spreading viral variant 1. This is because the second viral variant is 100-fold less common at the beginning of the simulation. Thus, with sufficient cross-protection (*ε*_12_ = *ε*_21_ = 0.7 in Figure 2), and a reasonably fast vaccine roll-out (100 days), it is best to target the more prevalent strain. However, the initial advantage gained by being more prevalent dissipates rapidly as the transmission rate of the second viral variant increases. Thus, when the second variant spreads 50% faster than the first, vaccination against the second strain is marginally better (see also Figure 1) and when the second variant spreads twice as fast as the first, vaccinating against the faster spreading second strain dramatically reduces total deaths, as well as peak infection rates.

**Fig. 2.**
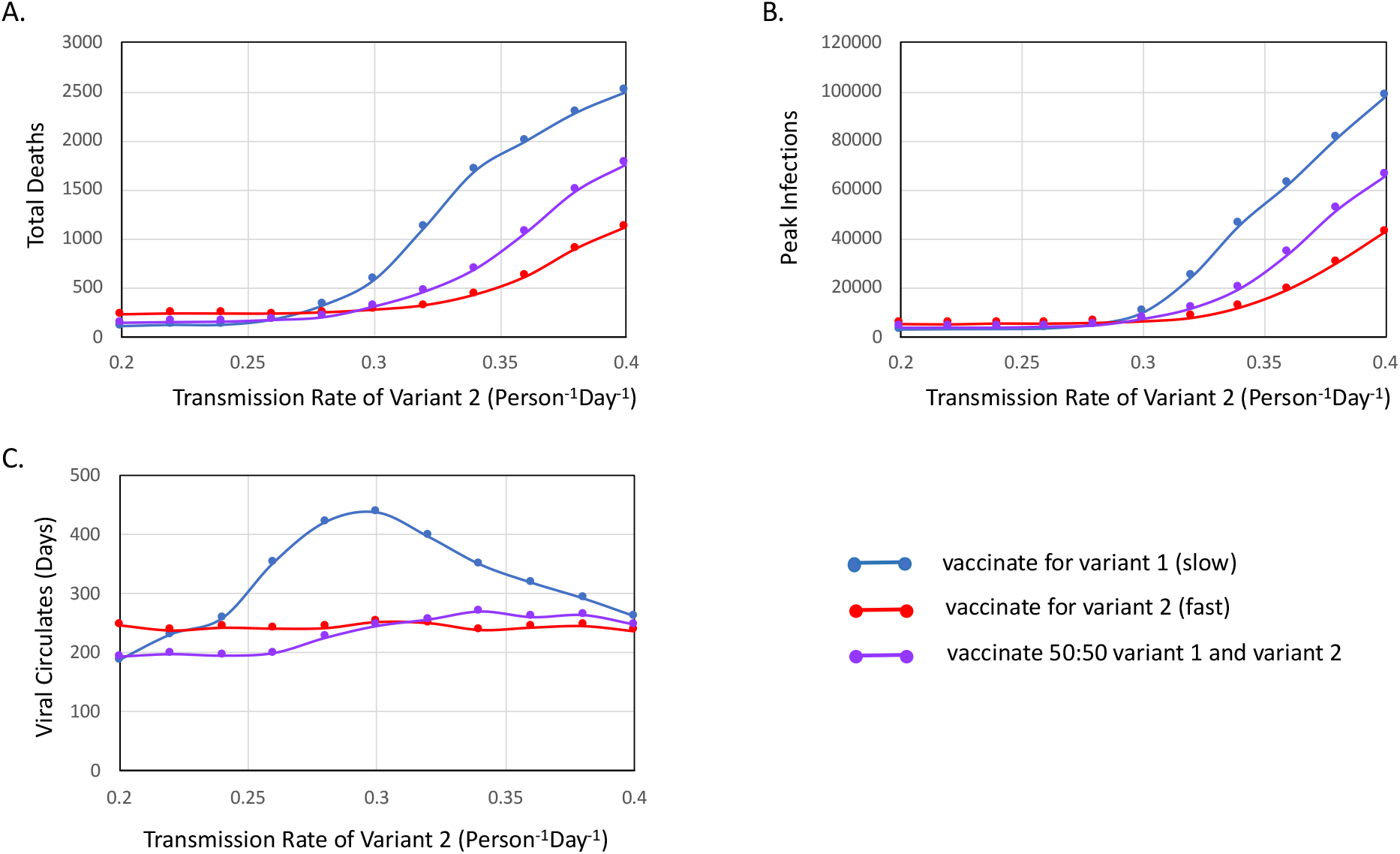
(A) Total number of deaths, (B) peak infections and (C) time to virus extinction as a function of the transmission rate of viral variant 2 (*β*_2_, faster spreading) assuming a vaccination strategy targeting viral variant 1 (blue, slower spreading), a vaccination strategy targeting viral variant 2 (red, faster spreading) and a mixed strategy with 50% of the population receiving the vaccine against viral variant 1 and the other 50% receiving the vaccine against viral variant 2. All parameters and initial conditions except the transmission rate of the second viral variant (*β*_2_) are as defined in Table 1. Results shown are median values over 30 trials.

For most of the range in Figure 2, the mixed vaccination strategy is intermediate to either single vaccination strategy in terms of total deaths and peak infections. However, while the difference between the mixed strategy and vaccination against viral variant 2 is minimal at *β*_2_ = 0.3 (see also Figure 1), this difference increases rapidly for *β*_2_ > 0.3, such that, when the second viral variant spreads twice as fast as the first, *β*_2_ = 0.4, vaccinating against the second variant save many lives, even compared to the mixed strategy. Thus, the benefits of the mixed strategy seen in Figure 1 do not necessarily hold when the second viral variant is significantly more transmissible. Notably, the benefit of developing vaccines against the faster spreading virus emerge despite the relatively high cross-protection levels (*ε*_12_ = *ε*_21_ = 0.7) assumed in Figure 2 and despite the 100-fold greater prevalence of the first variant at the beginning of the simulation.

Viral times to extinction are largely independent of the transmission rate of the second variant when the vaccine is targeted against this variant. This makes sense. When vaccination is against the second viral variant, the final stages of the outbreak are largely determined by spread of the first viral variant, meaning that the final stages of the outbreak are largely independent of the properties of the second viral strain. When the vaccine in use is against the first viral variant, however, the time to viral extinction is unimodal. This can be understood as follows: when the second viral variant spreads slowly, it can be rapidly brought under control by cross-protection from infection/vaccination against the first variant. However, rapid control through vaccination is less likely when the second variant spreads more rapidly. In this case, the second variant has greater potential to infect a larger fraction of the population before cross-protection, combined with natural immunity, finally bring the second variant under control. For faster and faster spread of the second variant, however, natural immunity occurs earlier and earlier, leading to a decrease in viral time to extinction.

### Vaccine Roll-out

Figure 3 shows the total number of deaths (A), peak infections (B) and time to virus extinction (C) as a function of the number of people vaccinated for the virus each day. For all of the simulations in Figure 3, I assume a population size of 500,000 individuals, thus it would take 100 days to vaccinate the entire population at a rate of 5000 people/day, and 50 days to vaccinate the entire population at a rate of 10000 people/day. As expected, total deaths and peak infection rates decrease sharply with increased rate of vaccination roll-out for all vaccination strategies.

**Fig. 3.**
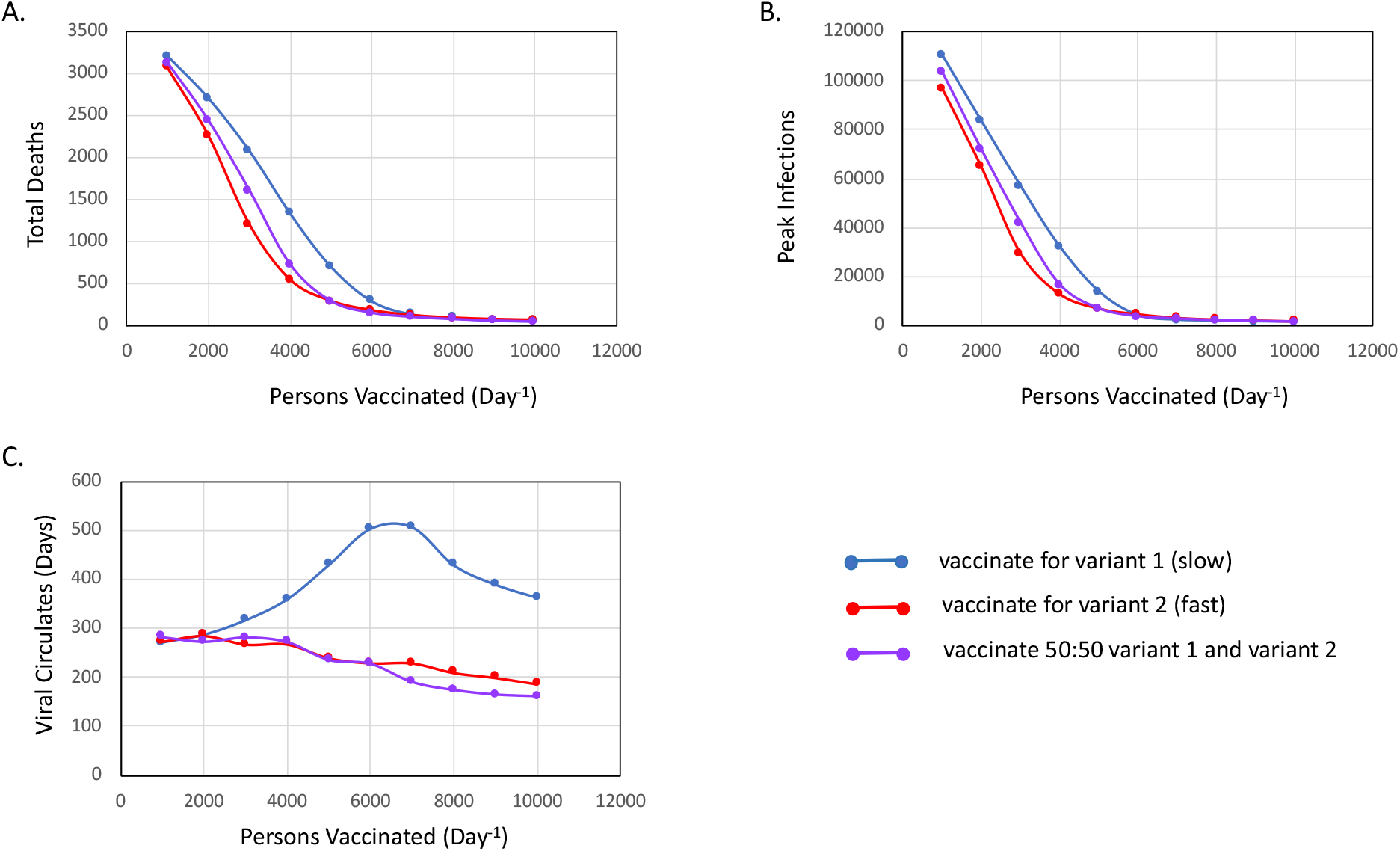
(A) Total number of deaths, (B) peak infections and (C) time to virus extinction as a function of vaccine roll-out rate, *ω*_1_ + *ω*_2_, assuming a vaccination strategy targeting viral variant 1 (blue, slower spreading), a vaccination strategy targeting viral variant 2 (red, faster spreading) and a mixed strategy with 50% of the population receiving the vaccine against viral variant 1 and the other 50% receiving the vaccine against viral variant 2. All parameters and initial conditions except overall vaccination rate (*ω*_1_ + *ω*_2_) are as defined in Table 1. Results shown are median values over 30 trials.

However, over the range from 1000-5000 people/day (i.e., 500-100 days to vaccinate the population), the decrease in deaths and peak infections as a function of vaccine rollout rate is greatest for vaccination against viral variant 2, intermediate for the mixed vaccination strategy and slowest for vaccination against viral variant 1. Thus, not surprisingly, slower vaccine roll-outs tend to favor vaccination strategies targeting the faster spreading viral variant. This is because slower vaccine roll-outs leave added time for the faster spreading variant to take over, even from a significant early deficit. When the vaccine roll-out is rapid (less than two months), however, all three strategies perform well. In fact, the mixed strategy outperforms both single vaccination strategies in terms of total deaths, and outperforms vaccination against variant 2 in terms of peak infections when the entire population can be vaccinated within 100 days. Further, when full vaccination can be completed within ∼60 days (2 months), vaccination against the first viral variant actually leads to the lowest peak infection rates, though it still gives higher total deaths than the mixed vaccination strategy.

For both vaccination against the fast-spreading virus and the mixed vaccination strategy, time to viral extinction decreases with increasing vaccine roll-out rates, commensurate with the expectation that faster vaccination of the population leads to faster suppression of the virus. Vaccination against the first viral variant, by contrast, leads to a peak in viral extinction times as a function of vaccine roll-out rate. Again, this is a result of the trade-off between rapid development of herd immunity at low roll-out rates, rapid development of vaccine-induced immunity at fast roll-out rates, and a peak in the middle, where vaccination slows viral spread down, but does not immediately suppress it.

### Initial Infection Rates

Figure 4 shows the total number of deaths (A), peak infections (B) and time to virus extinction (C) as a function of the number of people initially infected with viral variant 2. For these simulations, I keep the total number of initial infections constant, thus when there are more individuals infected with viral variant 2, there are fewer infected with viral variant 1. In order to span a wide range of variant 1: variant 2 infection ratios while maintaining at least 5 individuals infected with variant 2 (below 5 individuals, there is a sizeable likelihood of stochastic extinction of the second variant), I assume a much higher baseline infection rate in Figure 4 as compared to Figures 1-3. Specifically, in Figure 4, I assume that 1% of individuals are COVID-19 positive at the outset of the simulation, as compared to the 0.1% infection rate that I use in Figures 1-3.

**Fig. 4.**
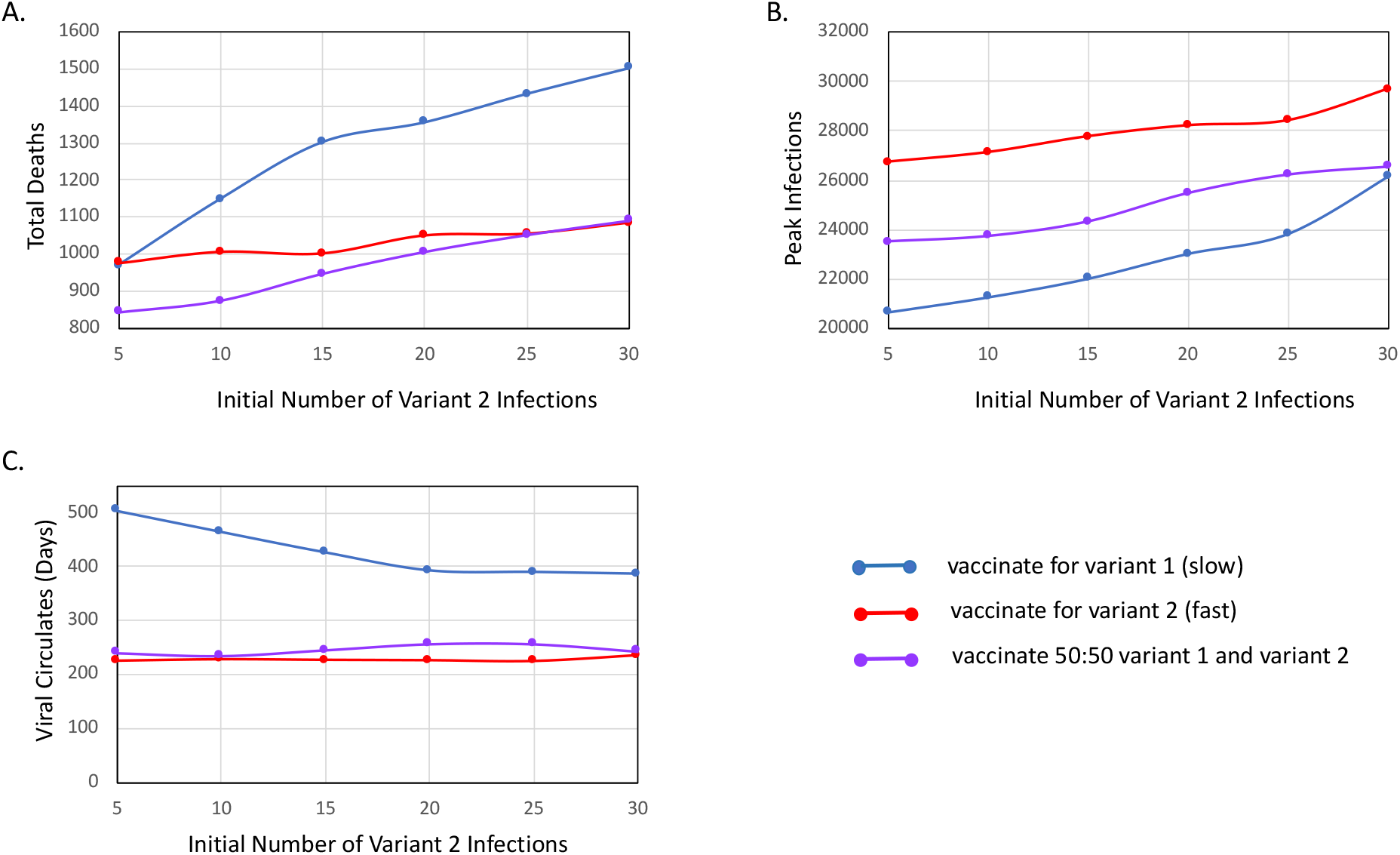
(A) Total number of deaths, (B) peak infections and (C) time to virus extinction as a function of the initial number of people in the population infected with viral variant 2, *ρ*_2_ ∑_*i,j,k*_ *N*, and assuming a vaccination strategy targeting viral variant 1 (blue, slower spreading), a vaccination strategy targeting viral variant 2 (red, faster spreading) and a mixed strategy with 50% of the population receiving the vaccine against viral variant 1 and the other 50% receiving the vaccine against viral variant 2. All parameters and initial conditions except initial fractions of infected individuals are as shown in Table 1. For initial fractions of infected individuals, I assume a constant total fraction, such that (*ρ*_1_ + *ρ*_2_) ∑_*i,j,k*_ *N* = 5005. Notice that this is a 10-fold higher initial infection rate than what is used in Figures 1-3. Results shown are median values over 30 trials.

**Fig. 4.**
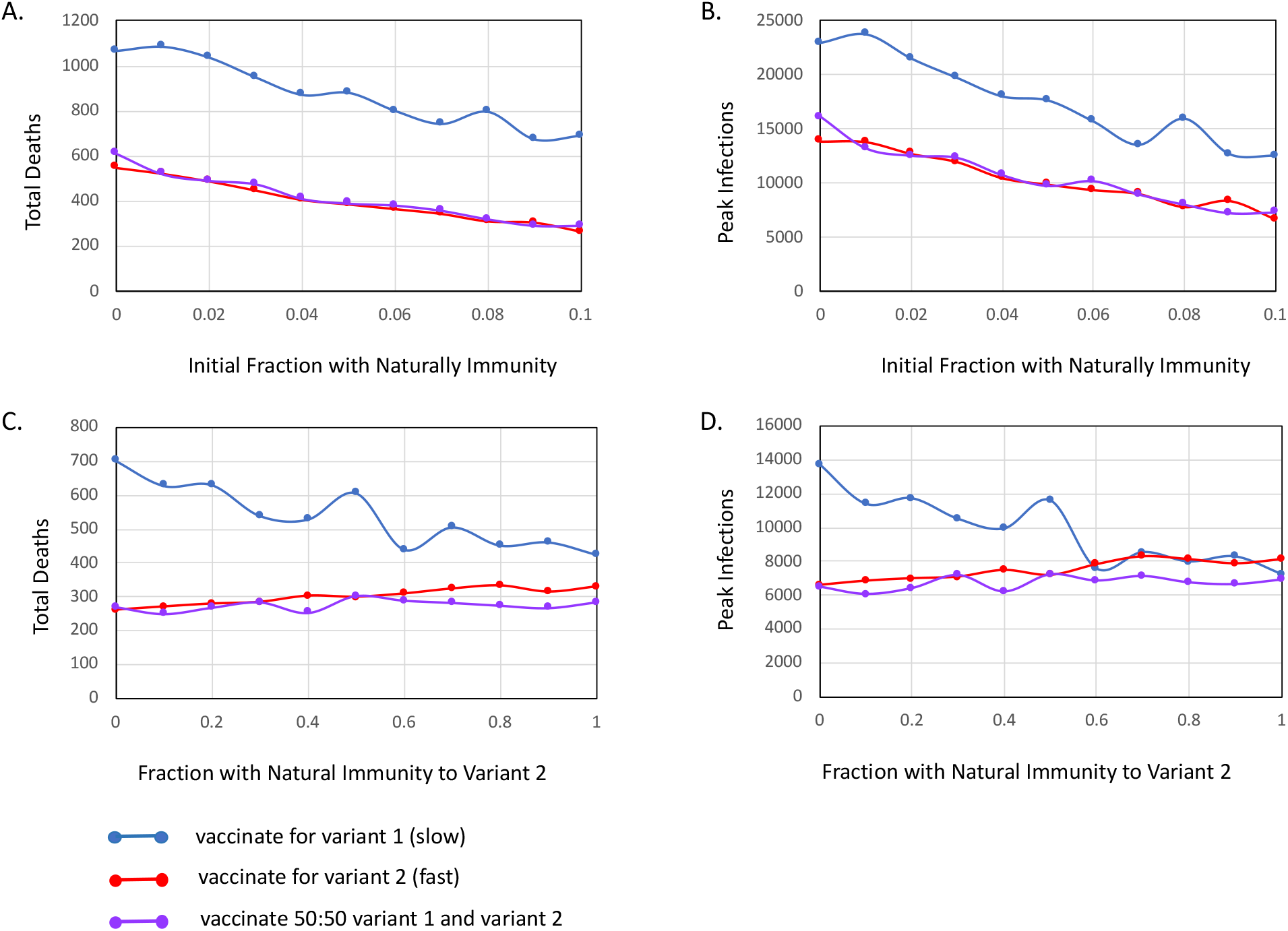
(A,C) Total number of deaths and (B,D) peak infections as a function of (A,B) the initial fraction of the population with natural immunity to viral variant 1 and assuming no immunity to viral variant 2 or (C,D) and the fraction of naturally immune individuals protected against viral variant 2, assuming an overall level of natural immunity of 10%. All parameters and initial conditions except initial fractions of naturally immune individuals are as shown in Table 1. Results shown are median values over 30 trials.

Despite the much higher prevalence of viral variant 1 in Figure 4, vaccinating against this variant is still generally less effective at preventing deaths. At best, vaccinating against the slow spreading viral variant 1 performs about as well as vaccinating against the fast spreading viral variant 2, and even this requires that the initial number of variant 2 infections be very low (≤ 5 total variant 2 infections and a 1000:1 ratio of variant 1 to variant 2 infections in a population of 500,000). More commonly, however, vaccinating against the slow spreading variant 1 leads to many more deaths than vaccinating against the fast spreading variant 2. While vaccinating against the slow-spreading strain is a poor strategy, the mixed vaccination strategy actually outperforms both single variant strategies over a wide range of infection ratios. Indeed, up to a variant 1:variant 2 infection ratio of approximately 200, the mixed strategy prevents the most deaths. Interestingly, although poor in terms of preventing deaths, vaccination against variant 1 can actually lower peak infection rates (but see Figures 1-3), suggesting that it is not necessarily a poor strategy along all public health dimensions. Initial infection ratios do not have strong impacts on time to virus extinction for any of the vaccine strategies considered.

### Existing Natural Immunity

Because COVID-19 has been spreading in most locations since February-March 2020, many people have acquired natural immunity, and this could potentially alter the benefits of the different vaccination strategies. I explore this effect in Figure 5. In Figures 5A,B, I assume that all existing natural immunity is towards the first viral variant, with the second viral variant being a very recent introduction. As expected, increasing overall rates of immunity to viral variant 1 at the start of the simulation decreases both total deaths and peak infection rates. Because, however, the rate of decrease is similar across all vaccination strategies, the overall level of natural immunity to the first viral variant has minimal impact on the choice of vaccination strategy. In Figures 5C,D, I consider a scenario where 10% of the population has natural immunity, but for some fraction of individuals this immunity is targeted against viral variant 1, and for others it is targeted against viral variant 2. Unlike changes in the overall levels of immunity, changes in the relative fraction of the population that is protected against each viral variant does impact performance of the different vaccinations strategies differently. In particular, when more of the population is already immune to viral variant 1, it accentuates the benefit of vaccinating against viral variant 2 and vice versa. This is because, when ∼10% of the population has natural immunity to viral variant 1(2), vaccinating against this variant means that ∼10% of vaccines are ‘wasted’. With decreasing levels of natural immunity against viral variant 1(2), however, the number of wasted variant 1(2) vaccines decreases. Interestingly, despite the loss of ‘wasted’ vaccines, vaccinating against the faster spreading second variant is still the better option for preventing deaths, and remains comparable for lowering peak infection rates even when all existing natural immunity is targeted against this strain. This shows how viral transmissibility swamps most other considerations when determining the optimal vaccine target. Notably, however, for all scenarios shown in Figure 5, vaccination against the second variant and the mixed vaccination strategy are comparable, suggesting that a mixed strategy can perform as well or almost as well as specifically targeting the fast-spreading strain (but see Figure 2).

## Discussion

In this paper, I use a stochastic simulation to examine how the choice of vaccine target impacts outcomes of an outbreak of COVID-19 consisting of two different viral variants and for which vaccine cross-protection is incomplete (though not necessarily zero). Although inspired by the recent emergence of the B.1.1.7 COVID-19 strain in the United Kingdom, predictions from this model are not restricted to any particular viral variant. Rather, model predictions hold for any virus strain that emerges and that exhibits a higher *R*0 than the dominant circulating strain – a typical trajectory for virus evolution. Indeed, as I write this paper, it is becoming clear that a second more transmissible COVID-19 strain has emerged in South Africa^23^. Like the UK B.1.1.7 strain, the South African variant has the N501Y mutation, although it is otherwise quite different.

As with increased transmission, viral variants that ‘escape’ current antibody responses are also anticipated as a common trajectory of viral evolution. Although many scientists are hopeful that existing vaccines will provide strong cross-protection against the B.1.1.7 viral variant from Southern England, the South African variant appears to have even more mutations in the spike protein. This reduces the likelihood that current vaccines will be able to neutralize it^24^. And even if current vaccines are largely effective against both the South England and South African viral variants, as more and more of the global population becomes immune to current circulating COVID-19 strains – either through natural infection or via vaccination – the selective pressures favoring escape mutants will increase. Thus, it is almost inevitable that fast-spreading variants that are not well-covered by existing vaccines will emerge during the course of vaccine roll-out across the globe.

One benefit of the technologies being used to develop COVID-19 vaccines and, in particular, of the mRNA vaccine approach, is the ease with which different viral variants can be exchanged during vaccine development and production. However, because mRNA vaccine technologies are relatively new, and have not been systematically tested in human populations prior to the COVID-19 pandemic, it is not clear whether *multiple* different viral targets can be included simultaneously (i.e., multivalent vaccines). Consequently, I sought to address the question of which viral variant to target in the event that a vaccine can only be produced against a single variant (monovalent) but there are multiple variants circulating.

Very broadly, my analysis suggests that it is almost always better to develop vaccines that target the faster spreading viral variant. This is true even when the slower spreading variant is 100- to 1000-fold more prevalent at the onset of the vaccination period and even when cross-protection is relatively high. This outcome is a direct result of the nature of exponential growth. In particular, exponential growth rapidly accentuates even slight differences in populations with different exponential growth rates (or different *R*0 values, in the case of diseases). As a consequence, over a matter of days to months, any initial advantage of the more prevalent but slower spreading viral variant is rapidly swamped by the diverging trajectories of the viral growth curves. Surprisingly, while targeting the slow spreading variant is rarely beneficial, a mixed strategy, where 50% of the population receives a vaccine against one strain, and 50% receives a vaccine against the other strain, can perform relatively well, at least when the differences in viral transmission rates are not extreme (e.g. the second viral variant is ≤50% more transmissible than the first). That said, for more extreme differences in transmission rates, the mixed strategy rapidly loses traction, and vaccination against the fast-spreading viral variant can save numerous lives, as well as lower peak infection rates, even relative to the mixed strategy.

While most realistic scenarios that I consider suggest that it is best to develop vaccines against the faster-spreading viral variant, this is not universally true. Factors that promote the viability of vaccination against the slow-spreading variant include smaller differences in growth rates between the two viral strains (see Figure 2), as well as faster vaccine roll-outs that leave less time for the fast-spreading virus to overcome its initial lower prevalence (see Figure 3). In addition, vaccination against the slow spreading viral variant is better when a larger relative fraction of the population is already immune to the fast spreading viral variant, when a larger relative fraction of the population is infected with the slow spreading viral variant, and when cross-protection is more complete. Thus, it is not inconceivable that there would be locations where vaccination against the slow spreading strain is preferable. Depending on how many people in Southern England have already been infected with B.1.1.7, for example, it may be more prudent to vaccinate against the original strain. Nevertheless, this appears to be the exception, rather than the rule. More generally, it seems that, for a largely susceptible population, and given current low rates of vaccine disbursement and delivery around the globe, it would be prudent if at least some vaccine companies switched their current formulations to target the newly emerged, highly transmissible COVID-19 variants that have been discovered in England and South Africa.

Although I designed my model to capture a range of important effects that could impact the benefits of different vaccination strategies, my model does make a number of simplifying assumptions. First, and foremost, I do not consider the two-dose vaccination schedule that is currently in use for both the Pfizer-BioNTech vaccine and the Moderna vaccine. Rather, my model ignores priming and assumes that a single vaccine dose results in a person acquiring full immunity. This simplifying assumption is most appropriate if the first dose of vaccine either does not provide much immunity, or else provides nearly complete protection. Predictions will be less accurate, however, if the first dose provides partial immunity that is then increased by the second dose. Another simplifying assumption of my model is that the population is well-mixed and without any structure like age-classes or differences in behavior (e.g., ability to telecommute, willingness to wear masks) that might lead to differential interactions among groups or else different exposure rates or susceptibilities to exposures. As well, I do not consider an exposed class or waning immunity. Likewise, I do not consider differences in infectious period or disease outcomes that may be garnered by immunity, either to the infecting strain or to the off-target strain. In reality, though, it is likely that vaccination would, at the very least, lower death rates of an infection, even if it does not fully protect against infection itself. Another assumption that I make is that, other than transmission rates, both viral variants are largely identical. That is, they induce similar death rates, have similar recovery rates, and illicit similar degrees of cross-protection. Finally, I assume a closed population, with no new individuals or infections introduced. Many of these complexities could be added to my model, though most would require additional parameters which are currently unknown. Nevertheless, as more information becomes available about newly emerging strains, their relative transmissibilities and the degree of cross-protection, including additional model details and complexities will become more feasible.

Overall, my model suggests that, except in very rare instances, monovalent COVID-19 vaccines should target the fastest-spreading strain of the virus, regardless of how prevalent that strain is at the outset of the vaccination period, and regardless of the degree of cross-protection offered by either vaccines or natural immunity. For scenarios where targeting the slower-spreading strain is equivalent or even marginally better than targeting the faster-spreading strain, total deaths and peak infection rates tend to be low for all vaccination strategies. However, for scenarios where targeting the faster-spreading strain is best, differences in total deaths and peak infections can be substantial. Thus, even from a precautionary principle, the safest bet is to target the variant with the higher transmission rate. The mixed strategy – vaccinating half of the population against each viral variant – performs nearly as well as vaccinating against the fast spreading virus over a surprisingly large range of viral transmission rates. However, when the fast spreading virus is significantly more transmissible than the original strain, even the mixed strategy can result in a sizeable number of additional deaths as compared to vaccination solely against the fast spreading strain. Thus, for example, for the low estimate that the B.1.1.7 strain is 50% more transmissible than its predecessor, the mixed vaccination strategy is nearly as good as vaccination against B.1.1.7 alone. However, for the high estimate that the B.1.1.7 strain is 70% more transmissible, the mixed strategy dramatically underperforms, leading to nearly 70% more deaths as compared to a strategy where all vaccine efforts are focused on the B.1.1.7 strain.

New strains of COVID-19 will continue to emerge that are more transmissible than the current variants, and that escape or partially escape from the current vaccines. Although we cannot prevent this from happening, we can make decisions about vaccination strategies that minimize the negative health outcomes of such events. Naturally, the best long-term solution will be to develop multivalent mRNA vaccines that simultaneously protect against all dominant COVID-19 viral variants in circulation. Until that is possible, however, my study provides ‘rule-of-thumb’ guidance for public health officials and vaccine companies alike. Specifically, my study suggests that, in most cases, targeting vaccines against the fastest spreading viral variant will at worst perform equally well as other strategies and, at best, save many lives.

## Data Availability

Python code for model simulations is provided at:
https://github.com/bewicklab/COVID-Vaccination-Strategies

